# To what extent physical distancing and other COVID-19 preventive measures being implemented among people in Arba Minch town, Southern Ethiopia: exploring evidences for an urgent call for action?

**DOI:** 10.1101/2020.08.11.20173211

**Authors:** Mekuria Asnakew Asfaw, Tsegaye Yohannes, Chuchu Churko, Alemayehu Bekele, Teklu Wegayehu

## Abstract

**Background:** The number of confirmed Coronavirus disease 2019 (COVID-19) cases surge substantially in resource-poor settings within the fragile health system. Since there are no proven vaccine and treatment in place against the disease, controlling strategy mainly rely on preventive measures. However, data on the extent of implementing physical distancing and other preventive measures were under estimated. This study, therefore, investigated these gaps among people in Arba Minch town, southern Ethiopia.

**Methods:** We conducted a community based cross-sectional study in Arba Minch town; from 15-30 June 2020. Data were collected using interviewer administered questionnaire and checklist. Then, data were cleaned, coded and entered to EpiData version 4.4.2, and exported to SPSS version 20 for analysis.

**Results:** Of the total participants (459), 43.6% achieved above the mean score (6±1.97) on preventive measures of COVID-19. Only 29.8% of participants kept the recommended physical distance, and surprisingly, in all public gathering places the distance was not kept totally. In addition, of the total participants, only 37.7% had face-mask use practice; 20.5% had hand sanitizer use practice, and 13.1% were measuring their body temperature every two weeks. Moreover, 42.5% of participants avoided attendance in public gatherings; 44.7% stopped touching their nose, eye and mouth; 55.6% practiced stay-at-home; and 60% had frequent hand washing practice. Majority of participants (66.7%) practiced covering their mouth and nose while coughing or sneezing; 68.2% had treatment seeking behavior if they experience flue like symptoms; 69.1% practiced isolating themselves while having flue like symptoms; and 89.3% avoided hand shaking.

**Conclusions:** The findings of this study suggest that physical distancing and other COVID-19 preventive measures were inadequately implemented among people in Arba Minch town. Thus, an urgent call for action is demanding to mitigate the spread of the COVID-19 as early as possible before it brings a devastating impact.

## Introduction

The novel Coronavirus disease 2019 (COVID-19) is a serious infectious disease, caused by the Severe acute respiratory syndrome coronavirus 2 (SARS-Co V-2) [1, 2]. On 30 January 2020, the World Health Organization (WHO) Director General declared COVID-19 as public health emergency of international concern [3]. Human-to-human infection due to SARS-CoV-2 occurs mainly through air droplets, close contact with infected persons, particularly mucus membranes secretions from nose, mouth, or eyes, contaminated surfaces, and some studies suggest digestive tract transmission [4, 5]. Elder people and those with underlying medical conditions, such as cardiovascular disease, diabetes, chronic respiratory disease, and cancer have higher risk to develop serious illness and probably result in death [6-8].

Despite the level of advancement in health system, the daily World Health Organization (WHO) Coronavirus situation reports highlight the fast spread of COVID-19 across the United States, Europe and South East-Asia [9]. As of August 7, 2020, WHO report showed that the ongoing pandemic of SARS-CoV-2 infection has led 18, 614 177 cases (259 344 new) and 702, 642 deaths (6 488 new) globally in 215 countries. Of these, Africa accounts 848, 053 cases (13 906 new) and 15, 252 deaths (502) [9]. Although the number of confirmed novel COVID-19 cases reported in resource-poor settings is still relatively lower, recently it was noticed that cases substantially rise. However, there is a high likelihood the current number represents underestimates due to inadequate test accessibility. Thus, these conditions may change in the coming months [10].

In Ethiopia, the first case was reported in March 13, 2020, in 48-year old Japanese in Addis Ababa [11]. As of August 7, 2020, 20 336 cases (46) and 356 deaths (13 new) were reported [9].

The WHO advises people to implement different preventive measures against COVID-19 pandemic. According to the WHO recommendations, the best way to halt transmission of human-to-human is being well informed about the virus, how it spreads and implementing the preventive measures adequately [12]. In response to the COVID-19 pandemic, the WHO along with its partners has been leading global coordination to hold the spread and reduce devastating impact of the COVID-19 pandemic [13].

Since the first incidence of the virus in Ethiopia, the country has been implementing unprecedented measures to control the rapid spread of the ongoing COVID-19 [14, 15]. However, anecdotally, it has been observed that communities in setting with strong social interaction are neglecting physical distancing and other preventive measures of COVID-19. Moreover, to best of our knowledge, there are no published studies that assess implementation of preventive measures of COVID-19 among the general population in Ethiopia. This study, therefore, aimed to investigate the extent of physical distancing and other preventive measures among people in Arba Minch town, Southern Ethiopia to inform decision made on COVID-19.

## Methods

### Study setting

This study was conducted in Arba Minch town which is located at 505kms south of Addis Ababa, the capital of Ethiopia. The town has 11 *kebeles* (smallest unit of government administration/ Based on the 2007 census conducted by central statistical agency, the total projected population for 2020 is 120, 736 (60, 127 men and 60, 609 women) [16]. As any part of the country, the community members in Arba Minch town are at high risk for Coronavirus infection due to existence of strong social interaction in the society which could favour the virus transmission rapidly. Since the first incidence of COVID-19 cases in Ethiopia, quarantine and treatment center have been established in the town. Currently, few confirmed COVID-19 cases have been reported in Arba Minch town.

### Study design and period

A community based cross-sectional study was conducted to investigate the extent of physical distancing and other preventive measures among people in Arba Minch town; from 15-30 June 2020.

### Study population

The study population was head of household or any adult ≥18 years old in the selected households who were residents and available during the survey period. Individuals were excluded from the study in situation when they were seriously ill and unable to provide information.

### Sample size and sampling technique

The sample size was determined using single proportion formula, 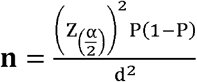, where, **p** is 50% (proportion of people implementing preventive measures), since there are no previous study conducted in the study area, **Zα/2** is the reliability coefficient of standard error at 5% level of significance = 1.96 and desired degree of precision **(d)** of **5 %**; the estimated sample size was **385**, and by adding **20%** non-response rate, the total computed sample size was 462. Study participants were selected by systematic random sampling technique from each *kebele* with consideration given to equal probability proportionate to sample size. For physical distancing measure, from each public gathering place, such as market, bank, church, *ekub*, hotels, bus station, and office distance between any two or more individuals was measured.

### Study variables

Variables included in this study were socio-demographic and economic characteristics, source of information, knowledge and perception on prevention and control of COVID-19; hygiene related factors, and implementation status of physical distancing, and other preventive measures.

### Data collection

Data were collected by house-to-house survey using interviewer administered questionnaire and observation checklist. Data quality was maintained by developing, adapting and pretesting standardized tool (adapted from WHO guidelines); training of data collectors and supervisors; and daily checking of consistency and accuracy of data.

### Statistical analysis

Data were edited, coded and entered to Epidata version 4.4.2 and exported to SPSS version 25 software. Then, the data were cleaned, analyzed and outcome of the research were presented by figures and tales. Implementing preventive measure was measured using 12 questions and score was computed by counting value within a case.

### Ethics statement

The study was reviewed and approved with reference number of IRB/412/12 by Institutional Research Ethics Review Board of College of Medicine and Health Sciences, Arba Minch University. Oral consent was received from sub-city administrators and heads of the household before data collection started. Data collectors and supervisors used face mask and alcohol based hand rub to safeguard them and participants while they collect data collect. In addition, they kept maintained physical distancing.

## Results

### Socio-demographic and economic characteristics

A total of 459 individuals participated in this study; giving a response rate of 99.4%. Table 1 presents detail on socio-demographic and economic data. The mean number of individual members in a household was 4.9±1.95. Of the total participants, more males participated than females (56.4% versus 43.6%). Almost 7% (32/459) of respondents earned less than 1000 Ethiopian Birr (ETB) per month, and 32.7% (150/459) did not have hand washing facility.

**Table 1.** Socio-demographic and economic characteristics of study participants in Arba Minch town, June, 2020.

| Characteristics | Category | Frequency | % |
| --- | --- | --- | --- |
| **Sex** | Female | 200 | 43.6 |
|  | Male | 259 | 56.4 |
| **Age category** | 18-29 | 86 | 18.7 |
|  | 30-39 | 123 | 26.8 |
|  | 40-49 | 126 | 27.5 |
|  | 50-59 | 66 | 14.4 |
|  | ≥ 60 | 58 | 12.6 |
| **Educational status** | Cannot read and write | 34 | 7.4 |
|  | Can read and write | 24 | 5.2 |
|  | Grade 1-8 | 61 | 13.3 |
|  | Grade 9-12 | 103 | 22.4 |
|  | College and above | 237 | 51.7 |
| **Occupation** | Farmer | 16 | 3.5 |
|  | Government employee | 169 | 36.8 |
|  | Business (self) | 136 | 29.6 |
|  | Unemployed | 33 | 7.2 |
|  | Others* | 105 | 22.9 |
| **Marital status** | Single | 42 | 9.2 |
|  | Married | 376 | 81.9 |
|  | Divorced | 22 | 4.8 |
|  | Windowed | 19 | 4.1 |
| **Number of household members** | <5 | 295 | 64.3 |
|  | ≥5 | 164 | 35.7 |
| **Monthly income (ETB)** | <1000 | 32 | 7.0 |
|  | 1000-2999 | 112 | 24.4 |
|  | 3000-4999 | 59 | 12.9 |
|  | 5000-5999 | 102 | 22.2 |
|  | 6000-7999 | 55 | 12.0 |
|  | 8000-9999 | 35 | 7.6 |
|  | ≥ 10000 | 64 | 13.9 |
| **Housing condition** | House or apartment with garden | 18 | 3.9 |
|  | Condominium | 10 | 2.2 |
|  | House or apartment in a building | 10 | 2.2 |
|  | House in a fence where many people live | 355 | 77.3 |
|  | Villa | 44 | 9.6 |
|  | Kebele house | 22 | 4.8 |
| **Obtain adequate water for hygiene** | Yes | 444 | 96.7 |
|  | No | 15 | 3.3 |
| **Hand wash facility** | Yes | 309 | 67.3 |
|  | No | 150 | 32.7 |
| **Soap available around hand wash facility (n=309)** | Yes | 281 | 90.9 |
|  | No | 28 | 9.1 |

\*=Housewife=51 and daily labourer= 54

### Source of information for COVID-19 pandemic

Of the total participants, 86.5% had access to COVID-19 related information from private television, and 60.4% were access from government television. The remaining respondents obtained information from social media, friends, radio, family members, and town crier (Figure 1).

**Figure 1.**
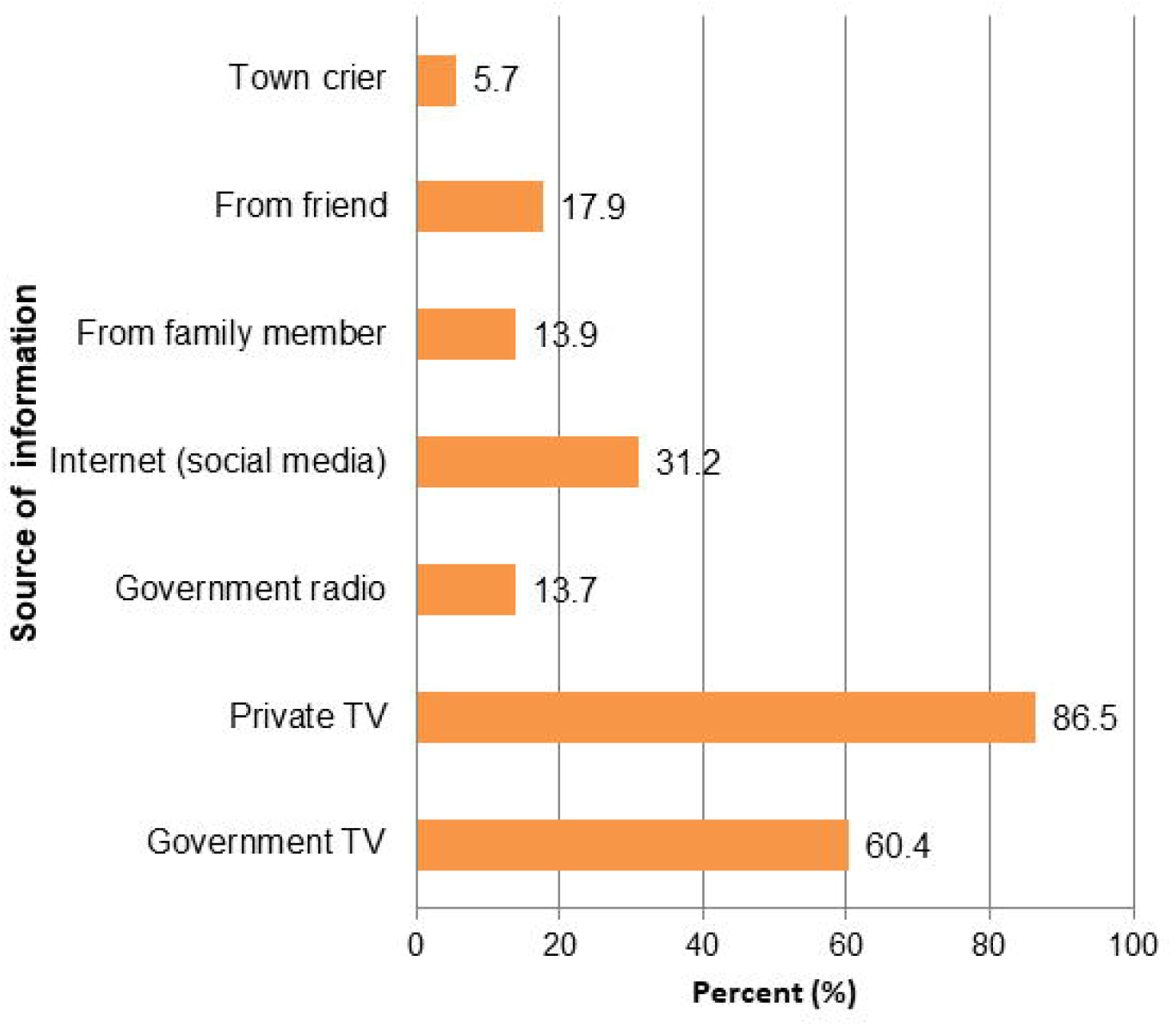
Sources of information related to COVID-19 pandemic in Arba Minch town, June, 2020.

### Knowledge, perception and other COVID-19 related information

Table 2 shows detail on awareness and COVID-19 related information among the study participants. Of the surveyed participants (459), almost all (99.3%) were informed on COVID-19. However, only 27.9% (128/459) responded that infected person are the main source of infection; 73.9% (339/459) knew COVID-19 symptoms, and 77.1% (354/459) believed that COVID-19 can be prevented. In addition, 3.7% (17/459) of participants faced psychological violence while implementing preventive measures; 6.5% (30/459) of respondents had history of in country travel in the last 7 days; and 14.4% (66/459) of participants worried about their health.

**Table 2.**
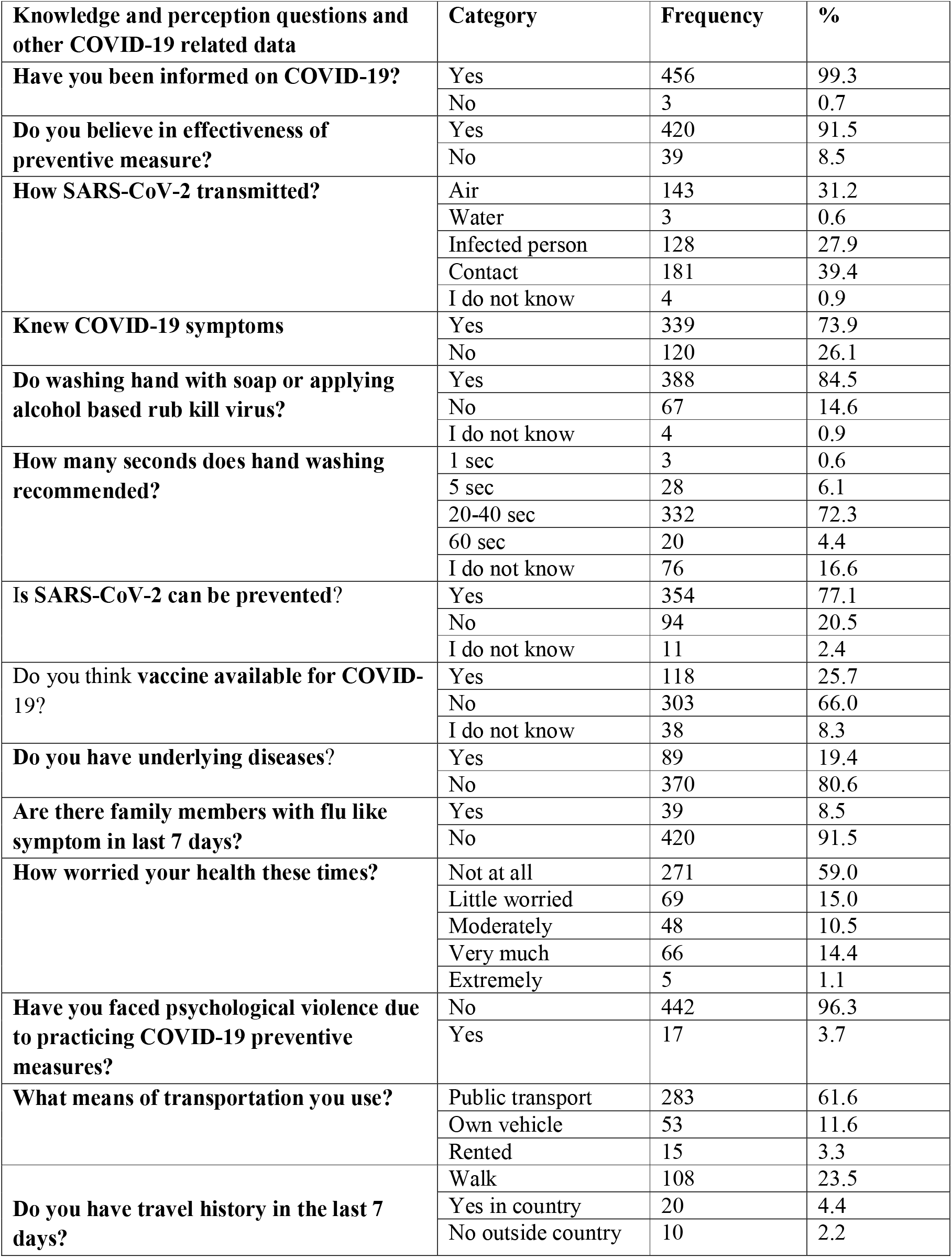
Knowledge, perception and other COVID-19 related information among study 206 participants, Arba Minch town, June, 2020 (n=459)

| **Knowledge and perception questions and other COVID-19 related data** | **Category** | **Frequency** | **%** |
| --- | --- | --- | --- |
| **Have you been informed on COVID-19?** | Yes | 456 | 99.3 |
|  | No | 3 | 0.7 |
| **Do you believe in effectiveness of preventive measure?** | Yes | 420 | 91.5 |
|  | No | 39 | 8.5 |
| **How SARS-CoV-2 transmitted?** | Air | 143 | 31.2 |
|  | Water | 3 | 0.6 |
|  | Infected person | 128 | 27.9 |
|  | Contact | 181 | 39.4 |
|  | I do not know | 4 | 0.9 |
| **Knew COVID-19 symptoms** | Yes | 339 | 73.9 |
|  | No | 120 | 26.1 |
| **Do washing hand with soap or applying alcohol based rub kill virus?** | Yes | 388 | 84.5 |
|  | No | 67 | 14.6 |
|  | I do not know | 4 | 0.9 |
| **How many seconds does hand washing recommended?** | 1 sec | 3 | 0.6 |
|  | 5 sec | 28 | 6.1 |
|  | 20-40 sec | 332 | 72.3 |
|  | 60 sec | 20 | 4.4 |
|  | I do not know | 76 | 16.6 |
| **Is SARS-CoV-2 can be prevented?** | Yes | 354 | 77.1 |
|  | No | 94 | 20.5 |
|  | I do not know | 11 | 2.4 |
| **Do you think vaccine available for COVID-19?** | Yes | 118 | 25.7 |
|  | No | 303 | 66.0 |
|  | I do not know | 38 | 8.3 |
| **Do you have underlying diseases?** | Yes | 89 | 19.4 |
|  | No | 370 | 80.6 |
| **Are there family members with flu like symptom in last 7 days?** | Yes | 39 | 8.5 |
|  | No | 420 | 91.5 |
| **How worried your health these times?** | Not at all | 271 | 59.0 |
|  | Little worried | 69 | 15.0 |
|  | Moderately | 48 | 10.5 |
|  | Very much | 66 | 14.4 |
|  | Extremely | 5 | 1.1 |
| **Have you faced psychological violence due to practicing COVID-19 preventive measures?** | No | 442 | 96.3 |
|  | Yes | 17 | 3.7 |
| **What means of transportation you use?** | Public transport | 283 | 61.6 |
|  | Own vehicle | 53 | 11.6 |
|  | Rented | 15 | 3.3 |
| **Do you have travel history in the last 7 days?** | Walk | 108 | 23.5 |
|  | Yes in country | 20 | 4.4 |
|  | No outside country | 10 | 2.2 |

### Status of implementing physical distancing

Of 55 surveyed public gathering places, the measured physical distances between any two or more people were less than 1 meter in 81.8% (45) of places. In addition, the recommended physical distance (at least 2 meters) was totally not kept in any of these places (Figure 2). On the other hands, of the total respondents (459)**, only 29.8**% (137) of participants self-reported as maintained at least 2 meter distance outside their home (Table 4).

**Figure 2.**
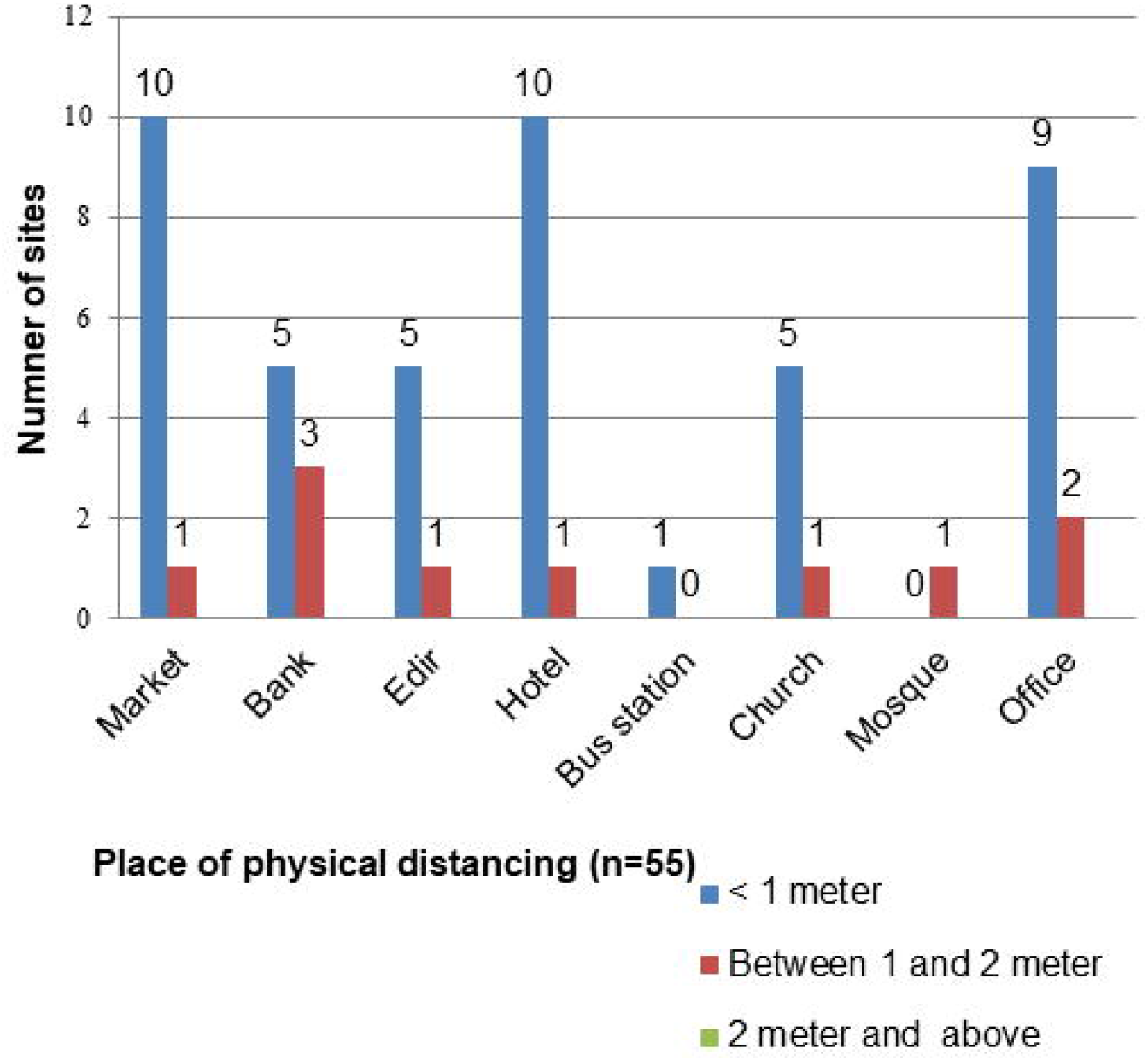
Physical distance between two or more individuals in different public gathering places of Arba Minch town, June, 2020 (n=55).

With regard to visiting crowded places, 54.9% (252/459) of participants visited market and 32.8% went to religious center (Churches and Mosques) in the last seven days (Figure 3).

**Figure 3.**
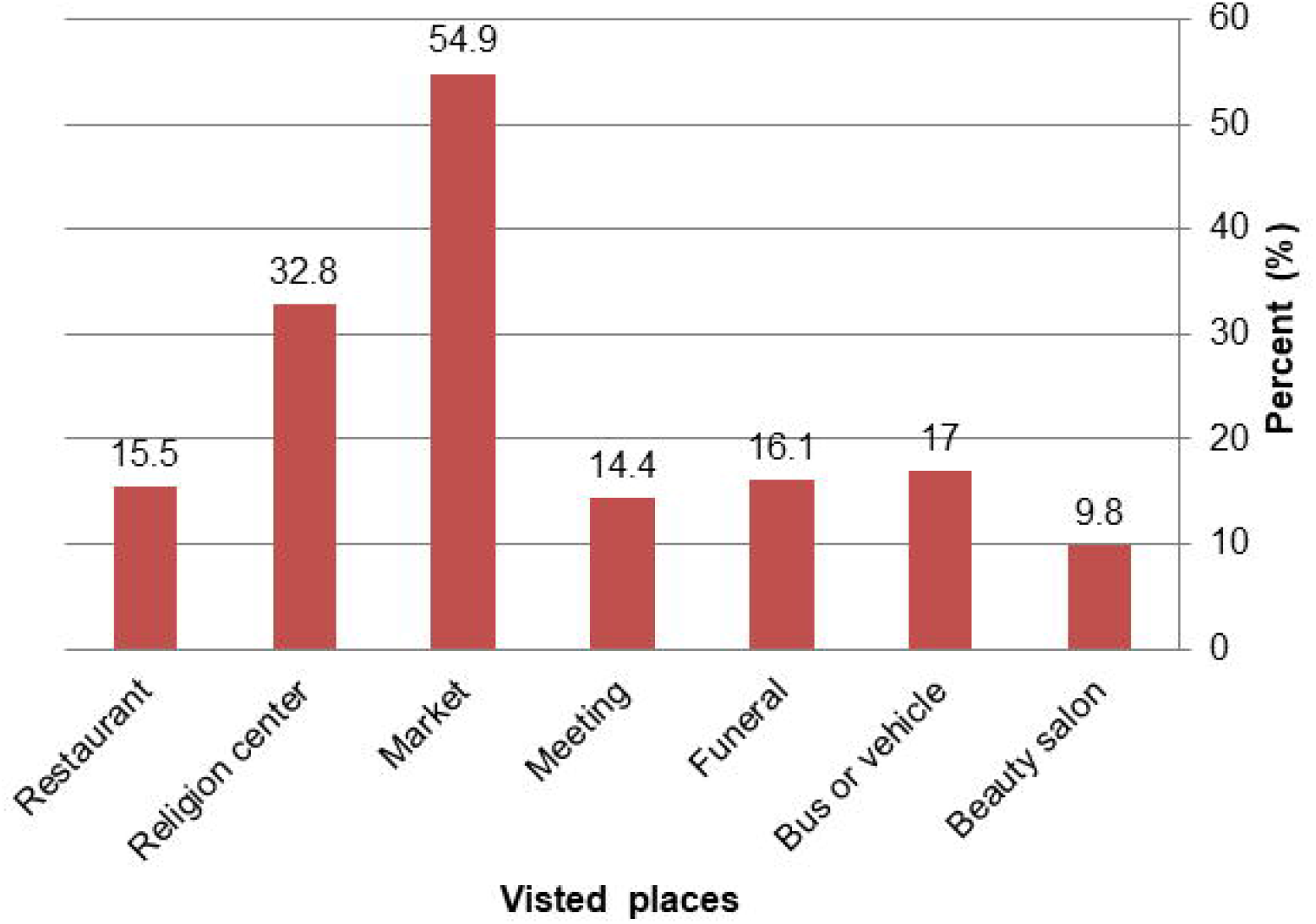
Public gathering places visited by participants in the last seven days in Arba Minch town, June, 2020.

### Status of implementing other preventive measures

We used 12 questions to assess implementation of preventive measures against COVID-19. In total, 43.6% (200/459) of participants achieved above the mean score (6±1.97) on preventive measures.

Of the surveyed individuals, only 37.7% (173/459) had face mask use practice, 20.5% (67/326) had frequent hand sanitizer use practice, and 13.1% (60/459) were measuring their body temperature every two week. Moreover, 42.5% (195/459) avoided going to public gathering place in the last 7 days; 44.7% (205/459) stopped touching their nose, eye and mouth; and 55.6 % (255/459) practiced stay-at-home if going outside is not mandatory; and 60% (254/423) had frequent hand washing practice. In addition, majority, 66.7% (306/459) practiced covering their mouth and nose while coughing or sneezing with cloth or tissue; 69.1% (317/459) practiced isolating themselves while having flue like symptoms; and mouth, 68.2% (313/459) had treatment seeking behavior if they experience flue like symptoms; and 89.3% (410/459) avoided hand shaking. Among those who did not use face mask, the main mentioned reason was not having money, 45.5% (130/286), to purchase the mask (Table 4).

**Table 4.**
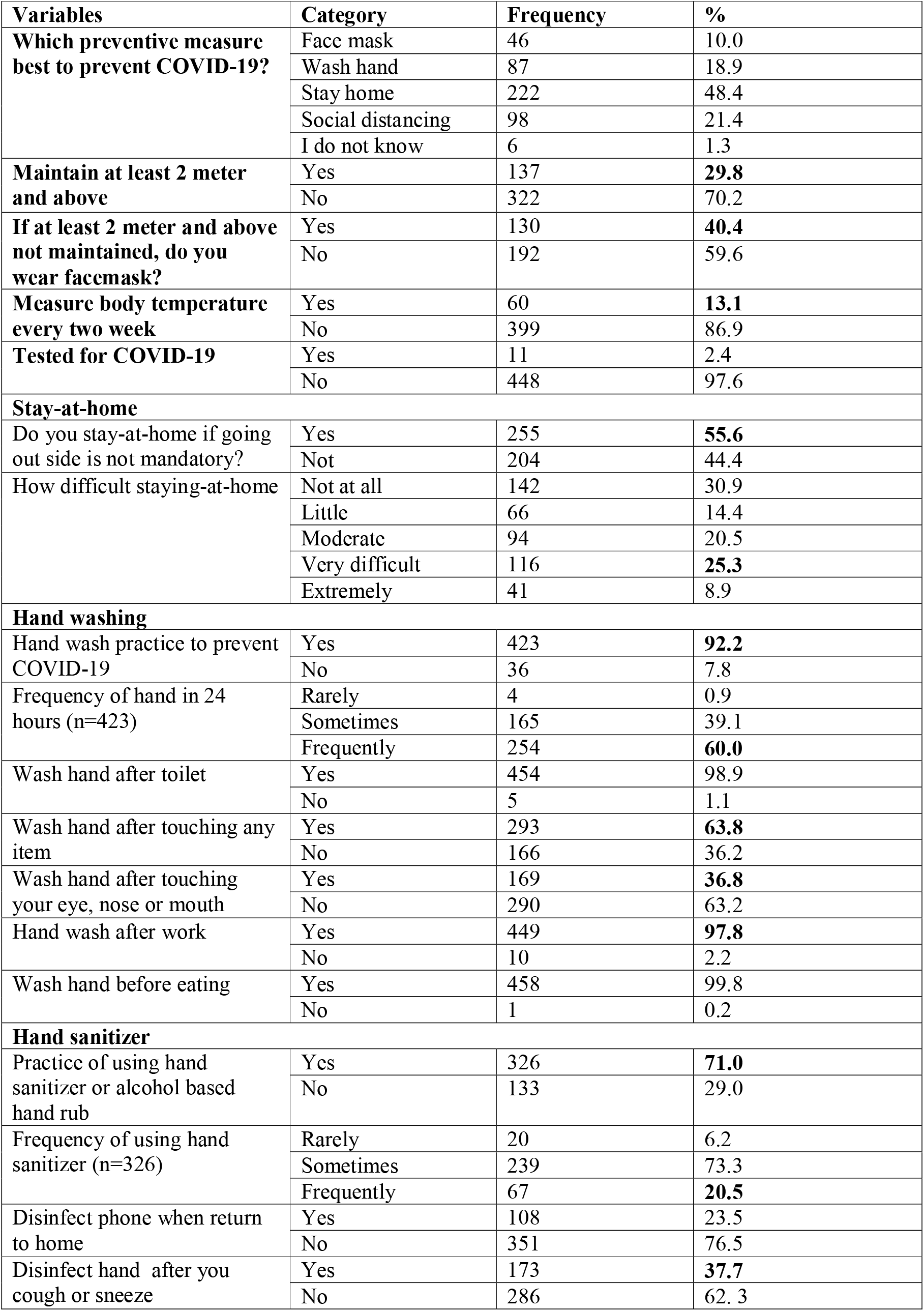
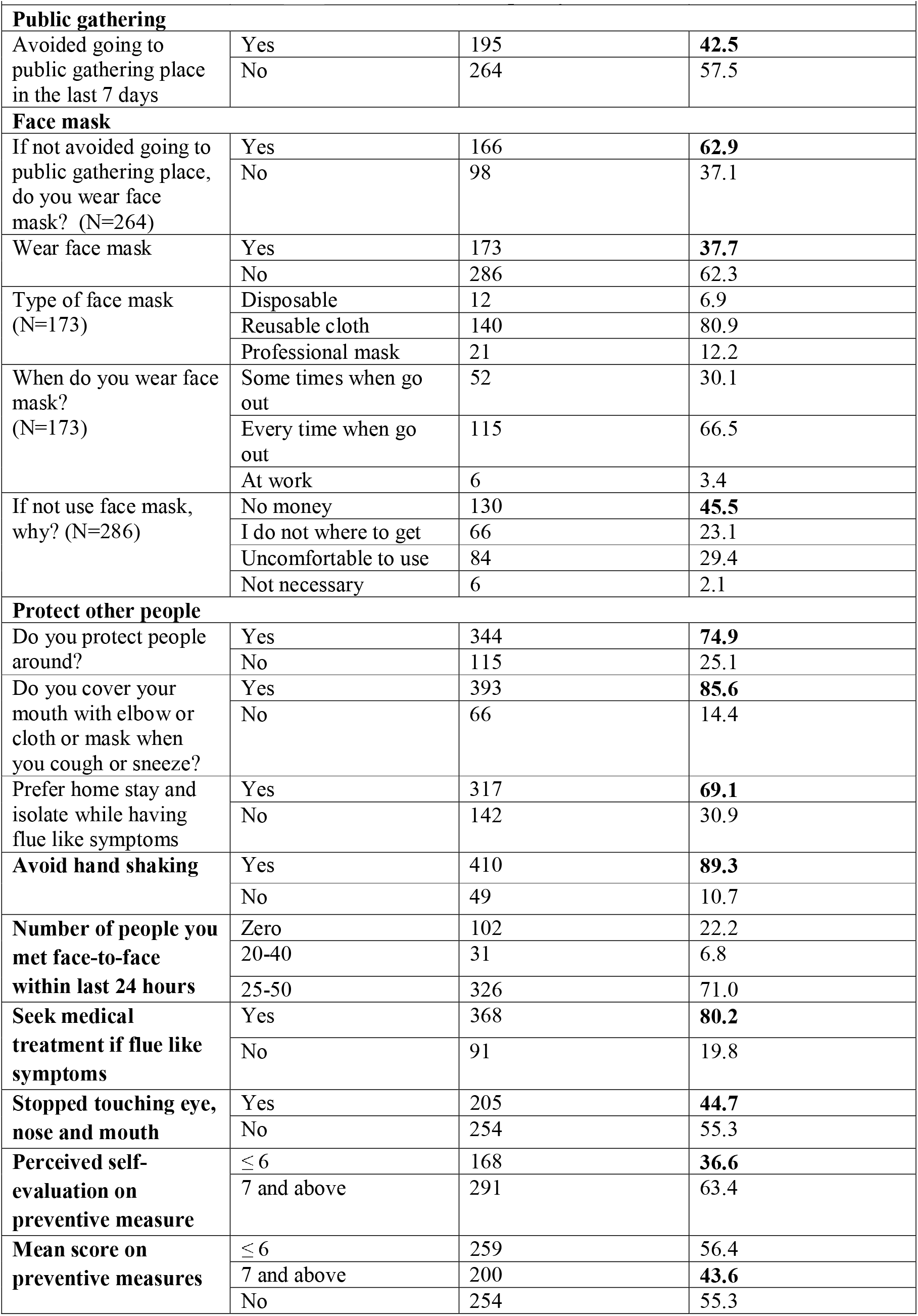
Implementing other preventive measures by study participants in Arba Minch town, June, 2020.

| Variables | Category | Frequency | % |
| --- | --- | --- | --- |
| **Which preventive measure best to prevent COVID-19?** | Face mask | 46 | 10.0 |
|  | Wash hand | 87 | 18.9 |
|  | Stay home | 222 | 48.4 |
|  | Social distancing | 98 | 21.4 |
|  | I do not know | 6 | 1.3 |
| **Maintain at least 2 meter and above** | Yes | 137 | **29.8** |
|  | No | 322 | 70.2 |
| **If at least 2 meter and above not maintained, do you wear facemask?** | Yes | 130 | **40.4** |
|  | No | 192 | 59.6 |
| **Measure body temperature every two week** | Yes | 60 | **13.1** |
|  | No | 399 | 86.9 |
| **Tested for COVID-19** | Yes | 11 | 2.4 |
|  | No | 448 | 97.6 |
| **Stay-at-home** |  |  |  |
| Do you stay-at-home if going out side is not mandatory? | Yes | 255 | **55.6** |
|  | Not | 204 | 44.4 |
| How difficult staying-at-home | Not at all | 142 | 30.9 |
|  | Little | 66 | 14.4 |
|  | Moderate | 94 | 20.5 |
|  | Very difficult | 116 | **25.3** |
|  | Extremely | 41 | 8.9 |
| **Hand washing** |  |  |  |
| Hand wash practice to prevent COVID-19 | Yes | 423 | **92.2** |
|  | No | 36 | 7.8 |
| Frequency of hand in 24 hours (n=423) | Rarely | 4 | 0.9 |
|  | Sometimes | 165 | 39.1 |
|  | Frequently | 254 | **60.0** |
| Wash hand after toilet | Yes | 454 | 98.9 |
|  | No | 5 | 1.1 |
| Wash hand after touching any item | Yes | 293 | **63.8** |
|  | No | 166 | 36.2 |
| Wash hand after touching your eye, nose or mouth | Yes | 169 | **36.8** |
|  | No | 290 | 63.2 |
| Hand wash after work | Yes | 449 | **97.8** |
|  | No | 10 | 2.2 |
| Wash hand before eating | Yes | 458 | 99.8 |
|  | No | 1 | 0.2 |
| **Hand sanitizer** |  |  |  |
| Practice of using hand sanitizer or alcohol based hand rub | Yes | 326 | **71.0** |
|  | No | 133 | 29.0 |
| Frequency of using hand sanitizer (n=326) | Rarely | 20 | 6.2 |
|  | Sometimes | 239 | 73.3 |
|  | Frequently | 67 | **20.5** |
| Disinfect phone when return to home | Yes | 108 | 23.5 |
|  | No | 351 | 76.5 |
| Disinfect hand after you cough or sneeze | Yes | 173 | **37.7** |
|  | No | 286 | 62.3 |
| **Public gathering** |  |  |  |
| Avoided going to public gathering place in the last 7 days | Yes | 195 | **42.5** |
|  | No | 264 | 57.5 |
| **Face mask** |  |  |  |
| If not avoided going to public gathering place, do you wear face mask? (N=264) | Yes | 166 | **62.9** |
|  | No | 98 | 37.1 |
| Wear face mask | Yes | 173 | **37.7** |
|  | No | 286 | 62.3 |
| Type of face mask (N=173) | Disposable | 12 | 6.9 |
|  | Reusable cloth | 140 | 80.9 |
|  | Professional mask | 21 | 12.2 |
| When do you wear face mask? (N=173) | Some times when go out | 52 | 30.1 |
|  | Every time when go out | 115 | 66.5 |
|  | At work | 6 | 3.4 |
| If not use face mask, why? (N=286) | No money | 130 | **45.5** |
|  | I do not where to get | 66 | 23.1 |
|  | Uncomfortable to use | 84 | 29.4 |
|  | Not necessary | 6 | 2.1 |
| **Protect other people** |  |  |  |
| Do you protect people around? | Yes | 344 | **74.9** |
|  | No | 115 | 25.1 |
| Do you cover your mouth with elbow or cloth or mask when you cough or sneeze? | Yes | 393 | **85.6** |
|  | No | 66 | 14.4 |
| Prefer home stay and isolate while having flue like symptoms | Yes | 317 | **69.1** |
|  | No | 142 | 30.9 |
| **Avoid hand shaking** | Yes | 410 | **89.3** |
|  | No | 49 | 10.7 |
| **Number of people you met face-to-face within last 24 hours** | Zero | 102 | 22.2 |
|  | 20-40 | 31 | 6.8 |
|  | 25-50 | 326 | 71.0 |
| **Seek medical treatment if flue like symptoms** | Yes | 368 | **80.2** |
|  | No | 91 | 19.8 |
| **Stopped touching eye, nose and mouth** | Yes | 205 | **44.7** |
|  | No | 254 | 55.3 |
| **Perceived self-evaluation on preventive measure** | $\leq 6$ | 168 | **36.6** |
|  | 7 and above | 291 | 63.4 |
| **Mean score on preventive measures** | $\leq 6$ | 259 | 56.4 |
|  | 7 and above | 200 | **43.6** |
|  | No | 254 | 55.3 |

### Difference in implementing preventive measures

Although numerical differences were noticed in implementing preventive measures by sociodemographic variables among participants, difference in availability of hand washing was statistically significant with hand washing practice at p-value <0.05 (Table 5).

**Table 5.** Socio-demographic characteristics related difference in implementing selected preventive measures by study participants toward COVID-19 (n = 459).

| Variables | Category | Preventive measures |  |  |  |  |  |
| --- | --- | --- | --- | --- | --- | --- | --- |
|  |  | Keep recommended physical distance |  |  | Use face mask |  |  |
|  |  | No | Yes | X<sup>2</sup> (P-value) | No | Yes | X<sup>2</sup> (P-value) |
|  |  | N (%) | N (%) |  | N (%) | N (%) |  |
| Sex | Male | 176 (68.0) | 83 (32.0) | **1.373 (0.241)** | 160 (61.8) | 99 (38.2) | **0.072 (0.788)** |
|  | Female | 146 (73.0) | 54 (27.0) |  | 126 (63.0) | 74 (37.0) |  |
| Age | 18-29 | 59 (68.6) | 27 (31.4) | **4.167 (0.384)** | 59 (68.6) | 27 (31.4) | **2.817 (0.589)** |
|  | 30-39 | 86 (69.9) | 37 (30.1) |  | 72 (58.5) | 51 (41.5) |  |
|  | 40-49 | 94 (74.6) | 32 (25.4) |  | 76 (60.3) | 50 (39.7) |  |
|  | 50-59 | 48 (72.7) | 18 (27.3) |  | 39 (59.1) | 27 (40.9) |  |
|  | ≥ 60 | 35 (60.3) | 23 (39.7) |  | 40 (69.0) | 18 (31.0) |  |
| Educational status | Cannot read and write | 27 (79.4) | 7 (20.6) | **2.836 (0.586)** | 26 (76.5) | 8 (23.5) | **8.928 (0.06)** |
|  | Can read and write | 15 (62.5) | 9 (37.5) |  | 17 (70.8) | 7 (29.2) |  |
|  | Grade 1-8 | 45 (73.8) | 16 (26.2) |  | 38 (62.3) | 23 (37.7) |  |
|  | Grade 9-12 | 73 (70.9) | 30 (29.1) |  | 71 (68.9) | 32 (31.1) |  |
|  | College and above | 162 (68.4) | 75 (31.6) |  | 134 (56.5) | 103 (43.5) |  |
| Occupation | Farmer | 13 (81.2) | 3 (18.8) | **5.243 (0.263)** | 13 (81.3) | 3 (18.8) | **2.817 (0.58)** |
|  | Government employee | 113 (66.9) | 56 (33.1) |  | 102 (60.4) | 67 (39.6) |  |
|  | Business (self) | 95 (69.9) | 41 (30.1) |  | 86 (63.2) | 50 (36.8) |  |
|  | Unemployed | 28 (84.8) | 5 (15.2) |  | 20 (60.6) | 13 (39.4) |  |
|  | Others* | 73 (69.5) | 32 (30.5) |  | 65 (61.9) | 40 (38.1) |  |

\*=Housewife=51 and daily labourer= 54

| Variable | Category | Hand wash regularly |  |  | Stay-at-home |  |  |
| --- | --- | --- | --- | --- | --- | --- | --- |
|  |  | No | Yes | X<sup>2</sup> | No | Yes | X<sup>2</sup> (P-value) |
|  |  | N (%) | N (%) | (P-value) | N (%) | N (%) |  |
| Sex | Male | 19<br>(7.3) | 240<br>(92.3) | **0.212**<br>**(0.646)** | 160<br>(61.8) | 99<br>(38.2) | 0.072 (0.788) |
|  | Female | 17<br>(8.5) | 183<br>(91.5) |  | 126<br>(63.0) | 74<br>(37.0) |  |
| Age | 18-29 | 10<br>(11.6) | 76<br>(88.4) | **5.024**<br>**(0.285)** | 59<br>(68.6) | 27<br>(31.4) | **3.795 (0.434)** |
|  | 30-39 | 12<br>(9.8) | 111<br>(90.2) |  | 72<br>(58.5) | 51<br>(41.5) |  |
|  | 40-49 | 6 (4.8) | 120<br>(95.2) |  | 76<br>(60.3) | 50<br>(39.7) |  |
|  | 50-59 | 3 (4.5) | 63<br>(95.5) |  | 39<br>(59.1) | 27<br>(40.9) |  |
|  | ≥ 60 | 5 (8.6) | 53<br>(91.4) |  | 40<br>(69.0) | 18<br>(31.0) |  |
| Educational status | Cannot read and write | 2 (5.9) | 32<br>(94.1) | **6.850**<br>**(0.144)** | 26<br>(76.5) | 8<br>(23.5) | **8.928**<br>**(0.063)** |
|  | Can read and write | 4<br>(16.7) | 20<br>(83.2) |  | 17<br>(70.8) | 7<br>(29.2) |  |
|  | Grade 1-8 | 1 (1.6) | 60<br>(98.4) |  | 38<br>(62.3) | 23<br>(37.7) |  |
|  | Grade 9-12 | 7 (6.8) | 96<br>(93.2) |  | 71<br>(68.9) | 32<br>(31.1) |  |
|  | College and above | 22<br>(9.3) | 215<br>(90.7) |  | 134<br>(56.5) | 103<br>(43.5) |  |
| Occupation | Farmer | 1 (6.3) | 15<br>(93.8) | **7.061**<br>**(0.133)** | 13<br>(81.3) | 3<br>(18.8) | **2.817 (0.589)** |
|  | Government employee | 19<br>(11.2) | 150<br>(88.8) |  | 102<br>(60.4) | 67<br>(39.6) |  |
|  | Business (self) | 5 (3.7) | 131<br>(96.3) |  | 86<br>(63.2) | 50<br>(36.8) |  |
|  | Unemployed | 4<br>(12.1) | 29<br>(87.9) |  | 20<br>(60.6) | 13<br>(39.4) |  |
|  | Others* | 7 (6.7) | 98<br>(93.3) |  | 65<br>(61.9) | 40<br>(38.1) |  |
| Hand washing facility | Yes | 18<br>(5.8) | 291<br>(94.2) | **5.327**<br>**(0.021)\*** | - | - | - |
|  | No | 18 (12) | 132<br>(88) |  | - | - |  |

\* = P-value significant at <0.05

## Discussion

This study explores data on adherence of people towards the recommended preventive measures of COVID-19. As any part of the country, the community members in Arba Minch town are at high risk for Coronavirus infection due to existence of strong social interaction in the society which could favour the virus transmission rapidly in the community. However, the findings of this study suggest that physical distancing and other COVID-19 preventive measures were inadequately implemented among people in Arba Minch town, southern Ethiopia. While almost all participants (99.3%) were informed on COVID-19, our study found out that only 43.6% of participants achieved above the mean score (6±1.97) on preventive measures. On the contrary, data of our study revealed that 63.4% of participants perceived as they were implementing the preventive measures against COVID-19.

In the current study, only 29.8% of participants self-reported as they kept at least 2 meters distance outside their home, and in none of the public gathering places the recommended physical distance (at least 2 meter) was not kept totally. The possible reason for low implementation of physical distancing is probably due to the strong social interaction norms that exist in the society. In consistent with this finding, a facility based study conducted in another part of Ethiopia (Jimma) revealed that slightly higher practice of avoiding physical proximity (33.6%) [17]. The higher report of keeping physical distance in Jimma probably due to proximity was measured at a distance with minimum of 1 meter.

In this study, only 37.7% of participants had face mask use practice when leaving out home, which is lower than the face mask use practice in Malaysia (51.2%) [18]. The lower practice of face mask in our study might be due to lack of money to purchase face mask, as justified by data of our study. Surprisingly, study participants in China demonstrated “as high as 98% of respondents had face mask use practice” [19].

With regard to hand sanitizer use, this study showed that only 20.5% of respondents had frequent use of hand sanitizer. The reason behind for low utilization of hand sanitizer in our study might be lack of access to hand sanitizer at affordable cost.

In the current study, only 13.07% of participants were measuring their body temperature every two week. The low practice of measuring body temperature is probably due to lack of access to temperature screening service.

In our study, we observed that less than 50% (42.5%) of respondents avoided going to public gathering places in the last 7 days. This result might be due to the fact that strong social interaction norm exist in the society, and our data justified as many people move to market areas to purchase their groceries. In consistent with this finding, a study conducted in Jimma town, Ethiopia demonstrated that a higher avoidance of going to public gathering place (53.8%) [17]. In addition, the finding of a study conducted in Malaysia showed a significant higher difference in avoiding going to public gathering places (83.4%) [18].

With regard to stopping touching nose, eye and mouth practice, in our study, 44.7% of participants stopped touching their nose, eye and mouth. This finding indicated that still more intervention is required to bring behavioral change.

This study demonstrated that 55.6 % of participants practiced stay-at-home as preventive measure. However, data of our study showed that substantial number of participants mentioned that stay-at-home is very challenging as a result of economic problem, which force people going outside their home to look for their daily breads.

Finding of the current study showed that only 60% of participant had frequent hand washing practice. The inadequate hand washing practice observed in this study could be due to lack of sustainable social behavioral change communication (SBCC). In consistent with this result, findings of studies conducted in another part of Ethiopia, in Philippines and Malaysia revealed much better hand washing practice [17, 18, 20].

Data of our study revealed that 66.7% of respondents had practice of covering their mouth and nose while coughing or sneezing with cloth, mask or tissue. Inadequate mouth and nose covering while coughing or sneezing with cloth, mask or tissue observed in this study could be due to lack of sustainable social behavioral change communication (SBCC). In the current study, 68.2% of participants had treatment seeking behavior if they experience flue like symptoms. This might be due to people have high fear of the virus as it could result in death.

Moreover, 69.1% of participants practiced isolating themselves while having flue like symptoms. In consistent with this finding, a bi-national study conducted in Africa (Nigeria and Egypt) showed that “as many as 96% of study participants practiced self-isolation and social distancing” [21].

Furthermore, predominantly, this study demonstrated that as high as 89.3% of participants avoided hand shaking. On the contrary, a lower practice of hand shaking was observed in a study conducted at another part of Ethiopia (53.8%) [17].

The main strengths are that we could assess community’s adherence towards the recommended preventive measures of the COVID-19 pandemic at community level, which address an important national and global operational research priority.

While interpreting data presented in this study, the following limitations need to be considered. First, findings are relied on self-reported practices of participants. Second, people may report as they were implementing preventive measures due to social desirability.

## Conclusions

The findings of this study suggest that physical distancing and other COVID-19 preventive measures were inadequately implemented among people in Arba Minch town. Thus, an urgent call for action is demanding in order to combat this dangerous infectious virus as early as possible before it brings devastating impact. Further, studies focusing on barriers relate to implementation of preventive measures against COVID-19 should be explored.

## Data Availability

All relevant data are within the manuscript.

## Acknowledgements

Authors would like to thank study participants, data collectors, and administrative officials.

